# Autoantibodies Targeting Angiotensin Converting Enzyme 2 Are Prevalent and Not Induced by SARS-CoV-2 Infection

**DOI:** 10.1101/2024.11.18.24317488

**Authors:** Yannick Galipeau, Nicolas Castonguay, Pauline S. McCluskie, Mayra Trentin Sonoda, Alexa Keeshan, Erin Collins, Corey Arnold, Martin Pelchat, Kevin Burns, Curtis Cooper, Marc-André Langlois

## Abstract

Clinical outcomes resulting from SARS-CoV-2 infection vary widely, ranging from asymptomatic cases to the development of mild to severe respiratory illness, and in some instances, chronic lingering disease and mortality. The underlying biological mechanisms driving this wide spectrum of pathogenicity among certain individuals and demographics remain elusive. Autoantibodies have emerged as potential contributors to the severity of COVID-19. Although preliminary reports have suggested the induction of antibodies targeting Angiotensin-Converting Enzyme II (ACE2) post-infection, this assertion lacks confirmation in large-scale studies. In this study, our objective is to comprehensively characterize and quantify the prevalence and expression levels of autoantibodies directed against ACE2 in a sizable cohort (n = 434). Our findings reveal that ACE2-reactive IgM antibodies are the most prevalent, with an overall seroprevalence of 18.8%, followed by IgG at 10.3% and IgA at 6.3%. Longitudinal analysis of individuals with multiple blood draws showed stable ACE2 IgG and IgA levels over time. Upon stratifying individuals based on molecular testing for SARS-CoV-2 or serological evidence of past infection, no significant differences were observed between groups. Functional assessment of ACE2 autoantibodies demonstrated that they are non-neutralizing and failed to inhibit spike-ACE2 interaction or affect the enzymatic activity of ACE2. Our results highlight that ACE2 autoantibodies are prevalent in the general population and were not induced by SARS-CoV-2 infection in our cohort. Notably, we found no substantiated evidence supporting a direct role for ACE2 autoantibodies in SARS-CoV-2 pathogenesis.

**Lay Summary:** This study examined the natural presence and function of autoantibodies targeting ACE2, the receptor for SARS-CoV-2, to determine if they influence COVID-19 severity. Using a cohort of over 400 individuals, including those with prior SARS-CoV-2 infection, we assessed the prevalence of ACE2-reactive IgM, IgG, and IgA antibodies in the general population. ACE2-reactive IgM antibodies were most common, found in approximately 18.8% of participants, followed by IgG at 10.3% and IgA at 6.3%.

Longitudinal analysis showed stable levels of IgG and IgA, with fluctuations in IgM over time. Importantly, no significant difference in ACE2 antibody levels was observed between individuals with or without SARS-CoV-2 infection, suggesting these autoantibodies were not induced by COVID-19. Functional tests showed that these ACE2 autoantibodies did not inhibit the virus’s spike-ACE2 interaction or alter ACE2’s enzymatic activity, indicating they are non-neutralizing.

We conclude that ACE2 autoantibodies are commonly present in the general population, independent of SARS-CoV-2 exposure, and are unlikely to play a role in COVID-19 severity. Further research is required to explore any potential physiological or pathological significance of ACE2 autoantibodies.

## Introduction

SARS-CoV-2 has profoundly impacted our day-to-day lives, in part through socio-economic impacts and excess mortality but also through morbidity associated with post-COVID-19 conditions (i.e., Long-COVID, PASC, PCC)(1). Although the scientific and medical communities have been intensely focusing on understanding SARS-CoV-2 pathogenesis, biological factors and mechanisms driving the heterogeneity of disease severity remain unclear. Several studies have highlighted that autoimmunity could be a factor impacting SARS-CoV-2 infection severity and recovery (2–4).

Links between viral infection and autoimmunity have been extensively documented. For example, Epstein-Barr virus (EBV) infection was recently linked with a 32-fold increased risk of developing multiple sclerosis (MS). While no causal or mechanistic evidence exists, as of today, EBV is thought to be the leading driver of MS (5). While evidence of autoimmune disease development following SARS-CoV-2 infections is currently anecdotal, several studies have reported the high prevalence of autoantibodies in COVID-19 patients. These autoantibodies include antinuclear antibodies, anti-cytokines antibodies, anti-phospholipid autoantibodies and others (6–11). Whilst some autoantibodies may have an important role in homeostasis and are not associated with specific diseases (i.e., natural autoantibodies) (12), most autoantibodies have been studied in the context of autoimmune pathologies. Interestingly, one group has shown that anti-IFN antibodies are detected in individuals with severe COVID-19, whereas none of the mild or asymptomatic individuals had anti-IFN autoantibodies in circulation in their cohort. These autoantibodies were able to neutralize type 1 IFNs, preventing their antiviral functions (8), thereby suggesting a role for autoantibodies in SARS-CoV-2 pathogenesis. These autoantibodies may represent a physiologically significant mechanism for regulating aberrant or excessive cytokine responses (13–15).

ACE2’s role in the renin-angiotensin system pathway has been extensively described. Briefly, ACE catalyzes the formation of angiotensin-II from the removal of two amino acids from the precursor angiotensin-I. ACE2 further enzymatically removes the carboxy-terminal phenylalanine from angiotensin-II, yielding angiotensin-(1-7), with both having downstream effects that are important for maintaining blood pressure, electrolyte balance and fluid homeostasis. Of note, ACE2 can also generate angiotensin-(1-9) (16, 17). ACE2 is expressed across several organ systems from the intestinal tract, kidneys, heart, lungs, and several others (18). In addition, ACE2 has also been identified as the cognate receptor for three coronaviruses, SARS-CoV, SARS-CoV-2, and NL63 (19–21). Given the relevance of ACE2 in viral entry of SARS-CoV-2, some have suggested that autoantibodies targeting ACE2 could impact COVID-19 severity (3). In a small cohort, Arthur *et al.* reported that 81% of previously SARS-CoV-2 infected individuals expressed antibodies targeting ACE2. These autoantibodies were found in 93% of hospitalized individuals with SARS-CoV-2 but were undetected in the control cohort (22). In another cohort, Casciola *et al.* found an association between anti-ACE2 IgM autoantibodies and severe COVID-19, with the ability to activate the complement and impact the vascular endothelial environment (23).

While studies found corroborating evidence of ACE2 autoantibodies in individuals with COVID-19 (24, 25), others did not. For example, in a screen by Chang *et al.*, ACE2 autoantibodies were detected in only one individual of the study cohort despite the identification of numerous other autoantibodies in several other individuals (9). The implication of ACE2 autoantibodies in SARS-CoV-2 pathogenesis remains unclear as ACE2 autoantibodies have been detected and associated with other medical conditions (i.e., Parkinson’s disease, vasculopathy, rheumatoid arthritis and others) independently of prior SARS-CoV-2 infection (26–30). In addition, the limited size of the control cohorts in studies involving SARS-CoV-2 infected individuals impedes proper statistical interpretation of the results.

Here we performed an in-depth characterization of ACE2 autoantibodies (IgG, IgA, and IgM) in the sera of 464 individuals, including 131 individuals with a history of prior SARS-CoV-2 infection. In addition, we functionally characterized anti-ACE2 autoantibodies, notably whether they were able to inhibit ACE2 enzymatic activity or block ACE2 and SARS-CoV-2 spike interactions.

## Results

### Identification of previous SARS-CoV-2 infections in a cross-sectional cohort

To study ACE2 autoantibodies following SARS-CoV-2 infection, serum samples of individuals enrolled in Stop the Spread Ottawa study were used (31, 32). This community-based prospective cohort study on immune responses to SARS-CoV-2 infection and vaccination recruited individuals with a previous history of, or were at risk for, SARS-CoV-2 infection. Individuals with at least one blood draw by December 2021 with corresponding consent and demographic information were included in this study. In total, 464 individuals between ages 18-74 were included in this study with an average age of 43.5 years (Sd=13.5 years) among which 67.2% were female and the majority were Caucasian (89.7%). Within the cohort, 131 individuals were identified as having been infected by SARS-CoV-2 previously (Table 1, Supp. Fig.1 and 2). SARS-CoV-2 infections were either self-reported to the study coordinators following a positive PCR test, diagnosed by a physician, or identified by a positive result by rapid antigen testing.

**Table 1.** Participant demographics and health characteristics based on SARS-CoV-2 infection history. Individuals who reported SARS-CoV-2 infection whether following a positive PCR test or confirmation by a medical professional, and individuals who had serological markers of prior infection (anti-N & anti-S IgG) were classified as individuals with a previous history of SARS-CoV-2 infection.

|  |  | **Combined**<br>**(n= 464)** | **Previous history of SARS-**<br>**CoV-2 infection (n=131)** | **No history of SARS-CoV-2**<br>**infection (n=333)** |
| --- | --- | --- | --- | --- |
| **Age (y.)** |  |  |  |  |
|  | Mean (SD) | 43.5(13.5) | 46.5(15.4) | 42.3(12.5) |
|  | Median | 42.5 | 45.0 | 42.0 |
|  | Range | 18-74 | 21-74 | 18-74 |
| **Sex** |  |  |  |  |
|  | Female Nb.(%) | 312(67.2) | 75(57.3) | 237(71.2) |
| **Ethnicity Nb (%)** |  |  |  |  |
|  | White | 416(89.7) | 116(88.5) | 300(90.0) |
|  | Aboriginal | 8(1.7) | 1(0.8) | 7(2.1) |
|  | Arab | 9(1.9) | 2(1.5) | 7(2.1) |
|  | Black | 5(1.1) | 3(2.3) | 2(0.6) |
|  | South Asian | 5(1.1) | 3(2.3) | 2(0.6) |
|  | Other | 21(4.5) | 6(4.6) | 15(4.5) |
| **BMI** |  |  |  |  |
|  | Nb. with BMI Info | 222 | 58 | 164 |
|  | Mean (SD) | 27.8 | 28.5 | 27.6 |
|  | Median | 26.7 | 27.5 | 26.4 |
|  | Range | 17.6 - 53.2 | 21.8 - 41.5 | 17.6 - 53.2 |
| **SARS-CoV-2 diagnosis Nb**<br>**(%)** |  |  |  |  |
|  | Self-reported | 121(26.1) | 121(92.4) | - |
|  | Serology | 87(18.8) | 87(66.4) | - |
| **Vaccinated Nb (%)** |  | 19(4.1) | 4(3.1) | 15(4.5) |
| **ACE2 seroprevalence (%)** |  |  |  |  |
|  | IgG | 10.3 | 6.1 | 12.0 |
|  | IgA | 6.3 | 7.6 | 5.7 |
|  | IgM | 18.8 | 12.2 | 21.3 |
|  | IgG and/or IgA and/or IgM | 30.4 | 22.9 | 33.3 |

Additionally, any individuals with IgG antibodies against the nucleocapsid (N) and Spike (S) above seroprevalence positivity thresholds were also identified as having been infected by SARS-CoV-2. This allowed the inclusion of asymptomatic and unreported infections. Our seropositivity threshold based on N and S dual positivity, reported and used previously (32–34), was set at a 3% false discovery rate of the density distribution of a large set of pre-pandemic sera. A sensitivity analysis was also performed utilizing a more stringent definition of SARS-CoV-2 infection, only considering 88 individuals with serological evidence of previous SARS-CoV-2 infection (Supp. Fig. 3).

### Measurement and seroprevalence of ACE2 autoantibodies

IgG, IgA, and IgM antibodies able to bind recombinant ACE2 were measured using a chemiluminescent direct ELISA platform that has been used and validated extensively for populational-based serological studies (32, 33, 35–38). Sera dilution used to measure ACE2 antibodies was validated and optimized to minimize background signal. For setting a seropositivity threshold, a systematic approach as described before was used (39). To establish cut-offs for IgG and IgA, values over 2 standard deviations of the mean were excluded, the new mean was recalculated, and a final threshold was set at 2 standard deviations. IgM seropositivity threshold was similarly established by excluding any values over 2 standard deviations of the mean and by recalculating the new mean, but the final threshold was set at 1 standard deviation given the spread of IgM values. The cut-off thresholds were inspected for robustness (Figure 1). Any samples with a signal-to-threshold ratio of 1 or under were presumed negative. This allowed an evaluation of the relative seroprevalence of all main isotypes of antibodies able to recognize ACE2 in sera of a large adult cohort (Table 1). ACE2-IgM antibodies were the most common with a seroprevalence of 18.8%. IgA seroprevalence in the overall cohort was the lowest at around 6.3% followed by IgG at 10.3%. Only a small fraction of the individuals with ACE2 autoantibodies simultaneously had all three isotypes present. The most common combination was IgG and IgM being detected together in 7.8% of the individuals with ACE2 antibodies. Less than 3% of individuals with ACE2 autoantibodies had either IgG/IgA or IgA/IgM detected simultaneously (Figure 1).

**Figure 1.**
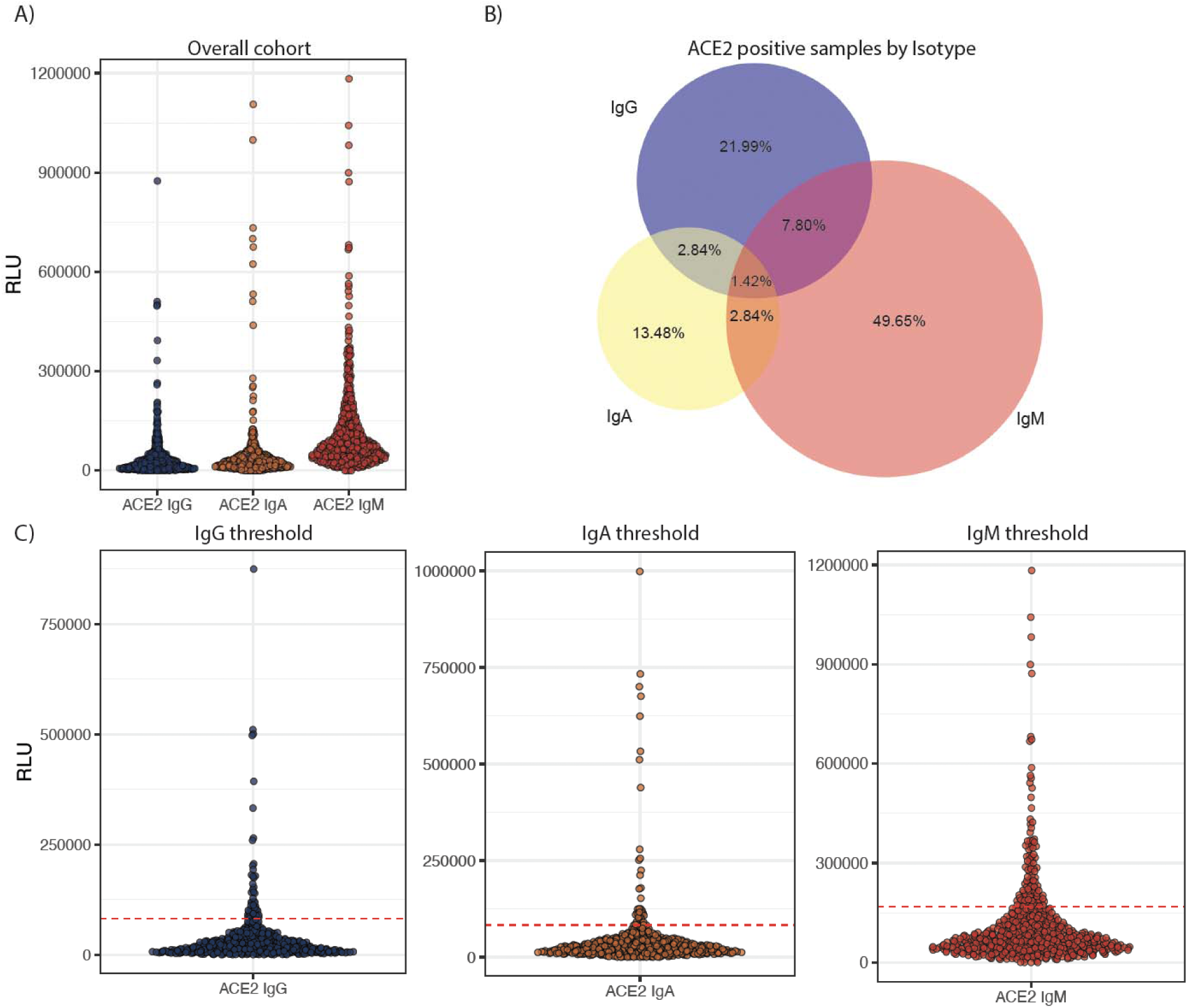
Overview of ACE2 serology results and establishment of seropositivity thresholds. A) overall view of anti-ACE2 IgG, IgA, and IgM antibodies of the full cohort including longitudinal sampling. B) Quantitative Venn diagram representation of the distribution between IgM, IgA, and IgG ACE2 autoantibodies in the overall cohort of ACE2-positive individuals. C) IgG, IgA, and IgM anti-ACE2 antibodies and associated threshold. IgG and IgA threshold was set at two cycles of 2 standard deviations (SD) from the mean of the presumed negative distribution. IgM threshold was set by a first cycle at 2SD and a second at 1SD from the mean of the presumed negative distribution.

### Clinical association and kinetics of ACE2 autoantibodies

Demographic and clinical data were available and matched to samples included in this study. The impact of several variables such as age, sex, co-morbidities, and diagnosed conditions on ACE2 autoantibody seroprevalence and levels was explored. No differences were seen for anti-ACE2 IgG and IgA between sexes, whereas levels of ACE2-IgM were elevated in the females in our cohort (p=0.00041) (Supp. Fig 4). For comparison of age, individuals were stratified into groups: ages 18-30, 31-40, 41-50, 51-60 and 60-74. No differences in the levels of anti-ACE2 IgG and IgA were observed between age groups. For anti-ACE2 IgM, individuals in the 18-30 group had elevated levels compared to the 41-50, 51-60, and 60-74 groups. Individuals in the 31-40 group also had higher levels when compared to the 60-74 group (Supp. Fig 5). While some age-related differences are statistically significant, the biological differences between groups are small. The association between seroprevalence of ACE2 autoantibodies seroprevalence and sex was assessed. Anti-ACE2 IgM seroprevalence was positively associated with female sex (OR 1.95 (1.12 - 3.38)) whereas males were associated with a lower seroprevalence of anti-ACE2 IgM (OR 0.51 (0.30 - 0.89)) (Figure 2, Supp. Table 1). A higher seroprevalence of anti-ACE2 IgG was observed for individuals with neurological conditions ((OR 5.48 (1.27 - 23.69)). This association was not seen with anti-ACE2 IgA or IgM. No other comorbidity or medical condition correlated with a higher or lower seroprevalence of ACE2 autoantibodies across all isotypes. During the study period, a subset of individuals provided one or two follow-up serum samples enabling us to investigate the kinetics of the various anti-ACE2 isotypes across time (Figure 3). Levels of IgG antibodies remained relatively stable over time (one individual displayed a strong increase at both subsequent sample collections). Interestingly, IgA levels remained relatively stable, with a few individuals showing a slight decay over time. As expected, anti-ACE2 IgM levels display a high degree of variability over time, likely due to the intrinsic half-life of IgM antibodies.

**Figure 2.**
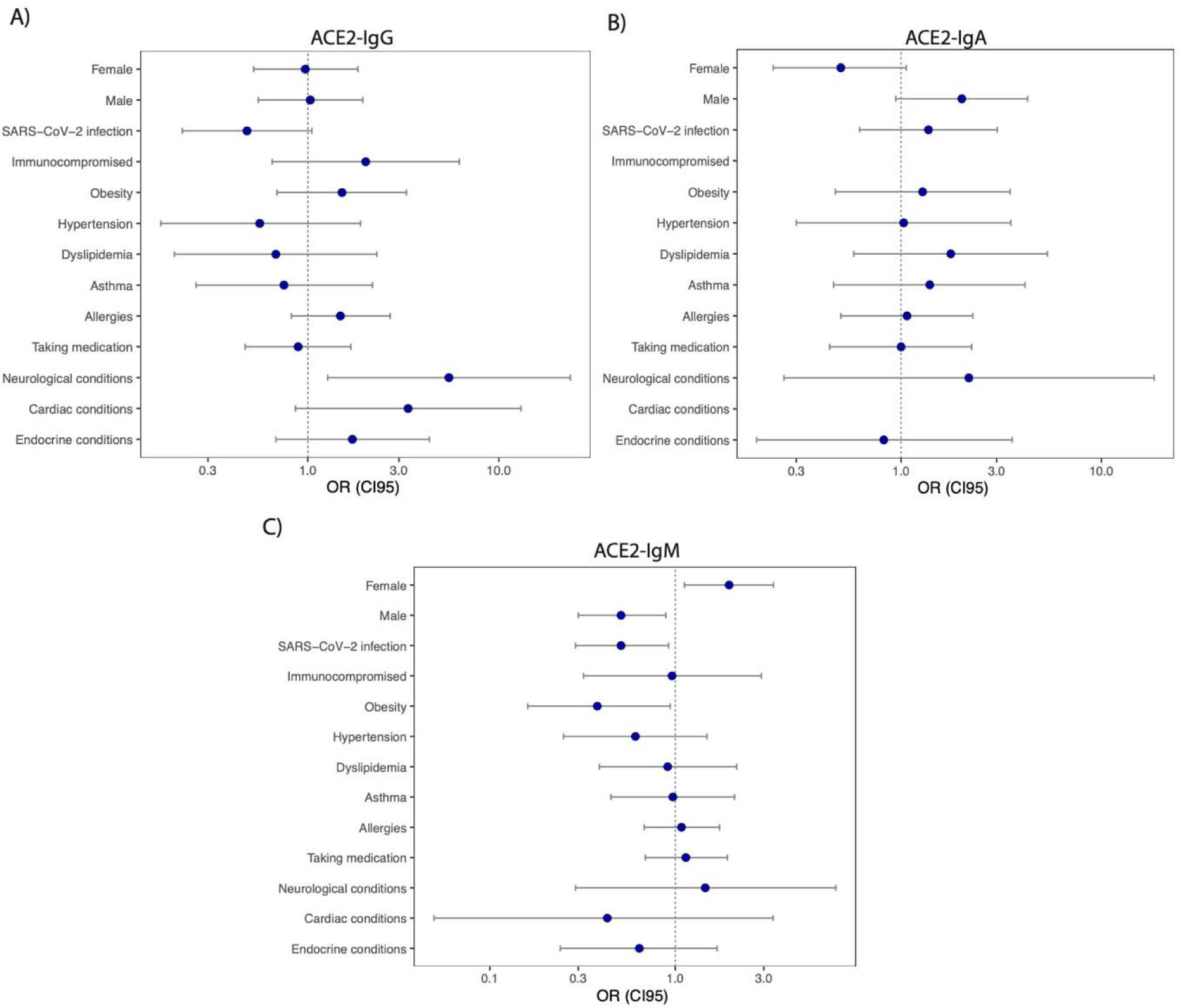
Influence of clinical features and demographic characteristics on ACE2 autoantibodie prevalence. Odd ratios (OR) and 95% confidence interval (CI) were calculated for demographi characteristics and clinical features for individuals in relation to A) IgG, B) IgA and C) IgM ACE2 autoantibodies. A table with a complete list of OR and 95% CI is available in the supplementary material (Supp. Table 1). The OR (CI95) are shown here on a log10 formatted axis.

**Figure 3.**
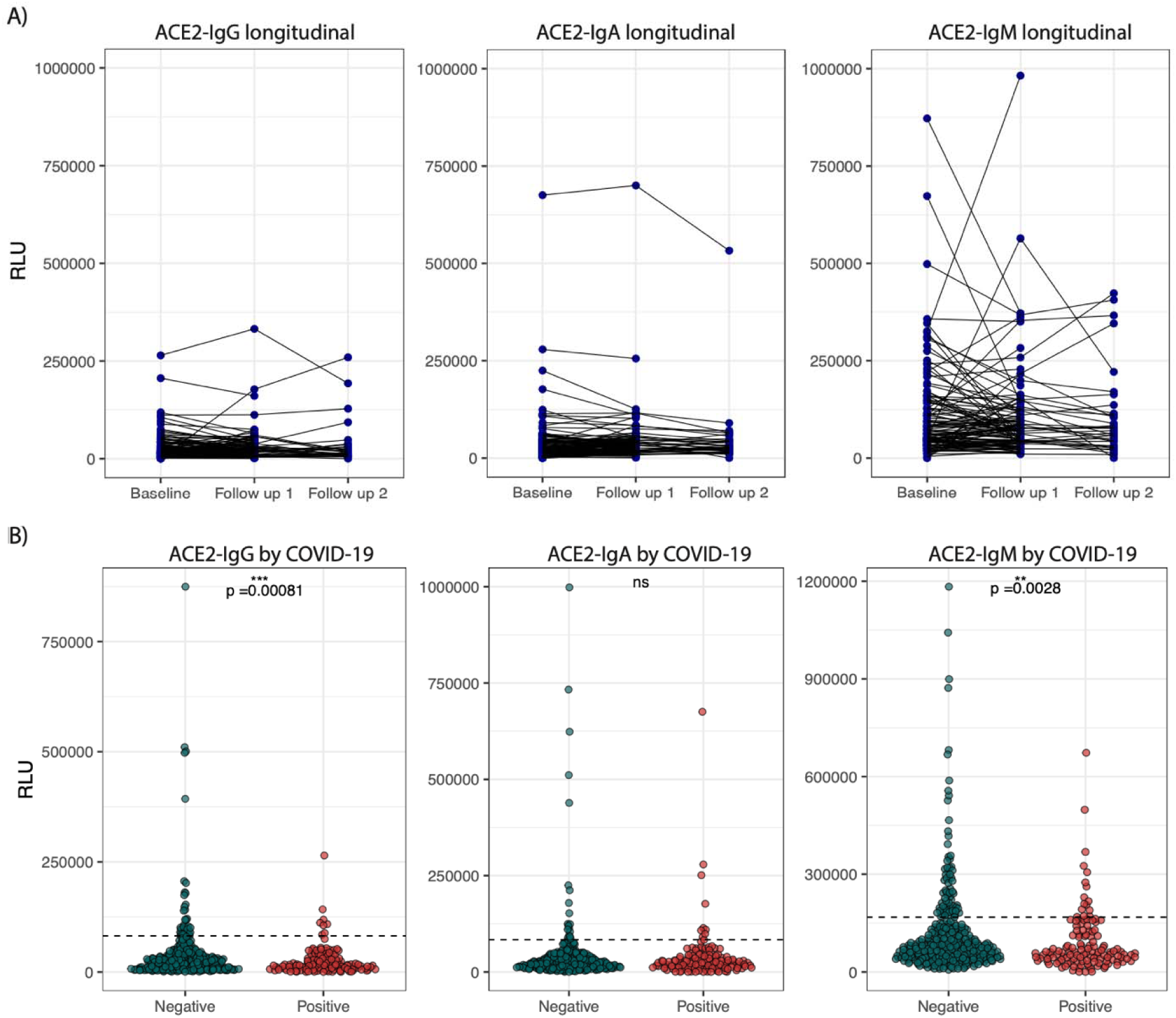
Persistence of autoantibodies in serum over time and association with previous SARS-CoV-2 infection. A) Some individuals had one or two follow-up samples. ACE2 IgG, IgA, IgM is shown longitudinally (Follow up 1 (n=120), median time between draws (SD) = 35 days (10), Follow up 2 (n=35), median time between draws (SD) = 37 days (6)). B) IgG, IgA, IgM ACE2 autoantibodies levels are shown for individuals who never had SARS-CoV-2 and for individuals who had serological markers of SARS-CoV-2 infection and/or self-reported a SARS-CoV-2 infection. Unpaired two-sided Wilcoxon test was used to establish statistical significance (n=464), * p< 0.05, ** p< 0.01, ***p<0.001, ****p<0.0001.

### Absence of association between previous SARS-CoV-2 infection and levels of ACE2 autoantibodies

Individuals in the overall cohort with previous SARS-CoV-2 infections were identified to investigate whether ACE2 autoantibodies are induced by COVID-19. To identify SARS-CoV-2 convalescent individuals, we employed 1) an inclusive definition of participants who self-reported a SARS-CoV-2 infection as well as participants with serological markers of previous SARS-CoV-2 infection, and 2) a stringent definition of participants that only had serological markers of previous infection. The levels of anti-ACE2 antibodies for each isotype were compared between individuals with no history of SARS-CoV-2 infection and individuals identified with previous SARS-CoV-2 infection using the inclusive definition (Figure 3) and individuals who only displayed serological markers of previous infection (Supp. Fig. 3). In both groups, no differences in anti-ACE2 IgG or IgA levels were detected. In both analysis groups, anti-ACE2 IgM antibodies were elevated in individuals with no history of previous infection in contrast to either convalescent group (p=0.006 & 0.0028). Likewise, prior history of SARS-CoV-2 was not associated with a higher or lower seroprevalence of IgA and IgG ACE2 autoantibodies. IgM ACE2 autoantibodies were associated with a lower seroprevalence in individuals with prior SARS-CoV-2 infections (OR 0.51 (0.29 - 0.92)) (Figure 2). Global analysis of ACE2 autoantibodies by PCA failed to identify convalescent individuals based on their levels of ACE2 autoantibodies (Supp. Fig 6). To investigate whether ACE2 autoantibody levels were associated with the intensity of humoral responses to recent SARS-CoV-2 infections we explored the association of IgG, IgA, and IgM ACE2 autoantibodies to the levels of IgG antibodies against SARS-CoV-2 nucleocapsid (Supp. Fig. 7). There was no association between ACE2 autoantibodies levels and nucleocapsid IgG intensity (R^2^ < 0.005). In this cohort, there is no evidence that prior SARS-CoV-2 infection results in a higher seroprevalence or levels of ACE2 autoantibodies in sera.

### ACE2 autoantibodies fail to inhibit ACE2 enzymatic activity or ACE2-spike interaction

While ACE2 autoantibodies are not elevated in levels or prevalence following SARS-CoV-2 infection in this cohort, their capacity to impact ACE2 either enzymatically or as a viral entry receptor was further investigated. A first subset of individuals (n=103) displaying variable levels of ACE2 autoantibodies were selected to investigate the ability of serum ACE2 autoantibodies to inhibit ACE2 enzymatic activity (Figure 4, Supp. Table 3). No differences in enzymatic activity were detected between sera positive for ACE2 autoantibodies (p=0.52) (Figure 4B). Interestingly, individuals who had a previous history of SARS-CoV-2 infection showed an elevated ACE2 enzymatic activity (p=0.00043) (Figure 4C). We next explored the potential of ACE2 autoantibodies to block SARS-CoV-2 spike and ACE2 interactions, as previous research has demonstrated the potential of using anti-ACE2 antibodies to block SARS-CoV-2 viral entry in vitro (40, 41). By modifying a previously developed protein-based surrogate neutralization assay (33), we measured the ability of sera from individuals with varying levels of ACE2 antibodies to inhibit ACE2/spike interaction (Figure 4D). Given that 65.5% of the samples were convalescent sera which contained antibodies against the spike protein (Supp. Table 3), these antibodies would confound our ability to measure the neutralizing contribution of ACE2 autoantibodies. As such, a series of depletion steps were performed by incubating sera in wells with recombinant SARS-CoV-2 spike protein to remove spike-reactive antibodies with potential ACE2-spike neutralizing capacity (Figure 4E). While this approach significantly reduced the levels of anti-spike antibodies, some samples retained relatively high levels of antibodies post-depletion, likely due to a high initial titer of anti-spike. The percent of inhibition of the interaction between SARS-CoV-2 spike and ACE2 was measured pre-and post-depletion (Figure 4F). A reduction in the ability of sera to inhibit ACE2/spike interactions was measured post-depletion confirming that anti-spike antibodies were a key source of neutralization. However, a few individuals displayed inhibition levels above 50% post-depletion. To understand if this was due to anti-ACE2 antibodies as opposed to anti-spike antibodies in samples where levels of anti-spike antibodies post-depletion remained elevated, post-depletion ACE2/spike inhibition levels were classified by ACE2 autoantibody seroprevalence (Figure 4G). No differences in inhibition were detected, regardless of whether or not samples had detectable levels of ACE2 autoantibodies. Rather, inhibition capacity of serum correlated with the levels of anti-spike antibodies post-depletion (Figure 4H). In addition, given that over 65% of the samples were from convalescent individuals, there is nothing to suggest that ACE2 autoantibodies from convalescent individuals differ (Supp. Table 3). The sera used in this assay was minimally diluted (1:1) with the assay buffer, with no neutralization detected. This contrasts SARS-CoV-2 antibodies which in our experience, on average can completely inhibit ACE2 and spike interaction at 1:25 to 1:125 in those same assay conditions. Overall, these results suggest that ACE2 autoantibodies at biological concentrations found in serum are unable to inhibit ACE2 and SARS-CoV-2 spike interaction.

**Figure 4.**
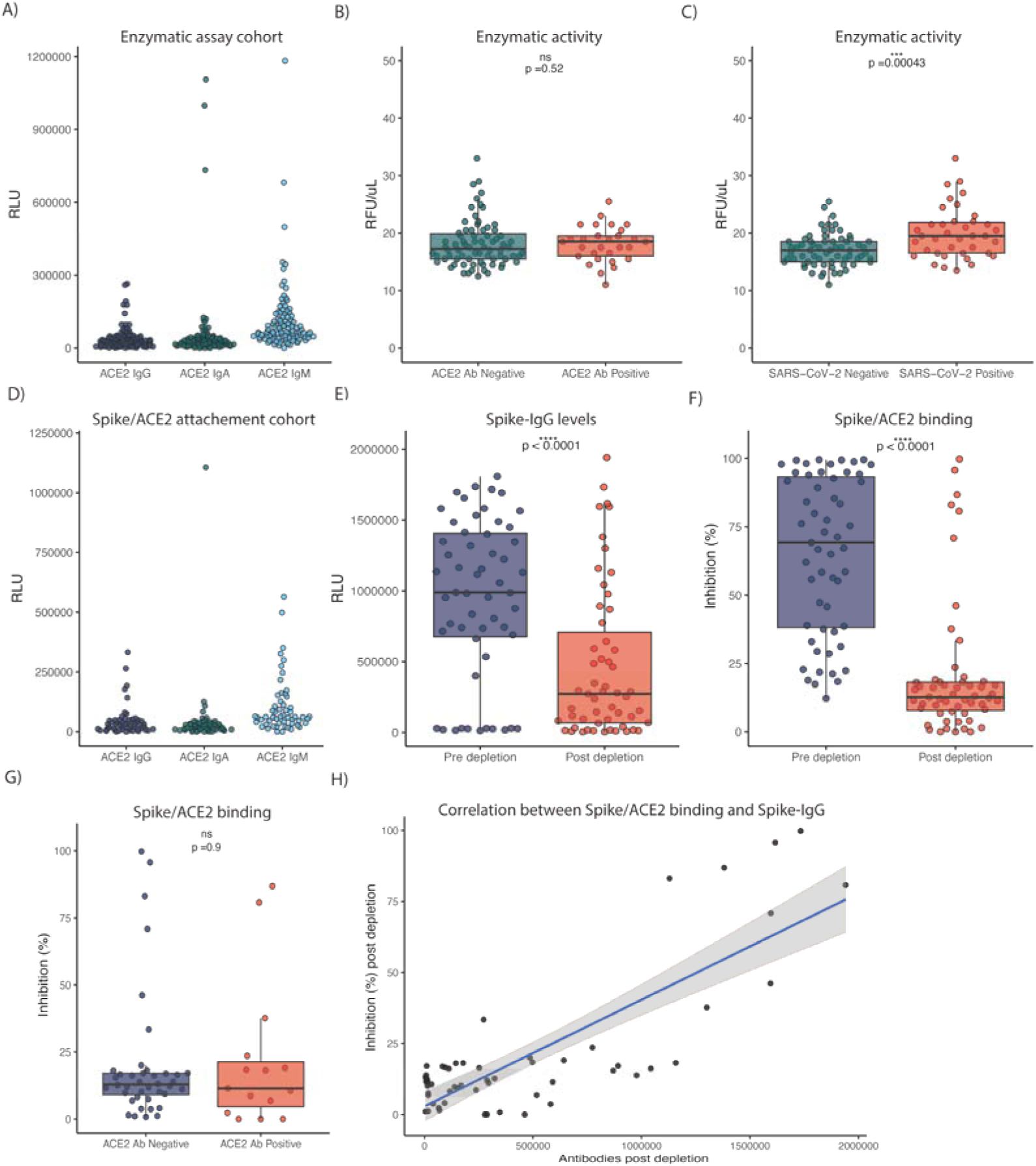
Effect of ACE2 autoantibodies on ACE2 enzymatic activity and SARS-CoV-2 spike-ACE2 binding. A) ACE2 autoantibodies levels (IgG, IgA, IgM) in a subset of samples for which ACE2 enzymatic activity was assessed. B) Levels of ACE2 enzymatic activity in individuals who had IgG and/or IgA and/or IgM ACE2 autoantibodies in contrast to the individuals who had no detectable ACE2 autoantibodies. C) Levels of ACE2 enzymatic activity in individuals who self-reported a SARS-CoV-2 infection and/or had serological markers of previous infection. D) ACE2 autoantibodies levels in a subset of samples in which ACE2 and spike interaction was assessed. E) Anti-Spike IgG antibodies levels pre-and post-depletion in sera. F) % inhibition representing the ability of the sera to inhibit Spike and ACE2 interaction pre-and post-depletion. G) % inhibition representing the ability of the sera to inhibit Spike and ACE2 interaction in individuals who had IgG and/or IgA and/or IgM ACE2 autoantibodies above threshold in contrast to the individuals who had no detectable ACE2 autoantibodies. H) Linear correlation between the ability of sera to inhibit Spike and ACE2 binding and the levels of Spike IgG antibodies in depleted sera. Unpaired two-sided Wilcoxon test was used to establish statistical significance where applicable, * p< 0.05, ** p< 0.01, ***p<0.001, ****p<0.0001.

## Discussion

ACE2 autoantibodies have been described in numerous studies prior to SARS-CoV-2 emergence (28). Previously, ACE2 autoantibodies have been linked with various health conditions such as Parkinson’s disease, systemic sclerosis, and rheumatoid arthritis (26, 27, 29). Given the role of ACE2 as the cognate receptor of SARS-CoV-2, scientific interest in ACE2 autoantibodies has resurfaced. Several studies have recently investigated ACE2 autoantibodies in small control and SARS-CoV-2 convalescent cohorts. These cohorts describing ACE2 autoantibodies have not simultaneously described the seroprevalence of all three main isotypes of antibodies (IgG, IgM, IgA) and the study sample sizes were small. By using the serum of individuals enrolled in the *Stop the Spread Ottawa Study* (31) we were able to generate a cross-sectional cohort of 333 healthy control individuals and 131 individuals with a laboratory-confirmed and self-reported history of previous SARS-CoV-2 infection thereby addressing sample size concerns (Table 1, Supp. Fig.1 and 2).

There are currently no recognized or standardized negative controls to establish seroprevalence to ACE2, and as such, the thresholds for this study were established by excluding high responders and setting the threshold from the presumed negative distribution of samples (Figure 1). We found that overall, nearly 1 in 5 individuals had detectable levels of ACE2-IgM. This echoes observations in a Japanese cohort that reported an ACE-IgM seroprevalence of 15% (30). For anti-ACE2 IgG antibodies, we estimated a seroprevalence of 10.3 %. A previous report of ACE2 IgG seroprevalence in a cohort of 20 control individuals was estimated at around 8.7% (29). Simultaneous IgM and IgG reactivity to ACE2 in individuals with detectable levels of ACE2 autoantibodies was estimated at 7.8% in our cohort, and a previous study had estimated this parameter at 6% (30). To our knowledge, we are the first group to measure IgA antibodies targeting ACE2. In our cohort, the seroprevalence of ACE2-IgA was 6.3%, with less than 3% of individuals simultaneously positive for ACE2 IgG, IgA and IgM autoantibodies. Overall, the percentage of individuals who had ACE2 autoantibodies of any isotype in our cohort was 30.4%.

Demographic information and co-morbidity data were collected alongside serum samples, which we leveraged to investigate how various factors may influence ACE2 autoantibody seroprevalence or their abundance intensity. Females had a higher seroprevalence and higher levels of ACE2-IgM. Stratifying by age, only significant differences in autoantibody levels were seen for ACE2-IgM, with older individuals displaying lower levels. Several types of autoantibodies including anti-nuclear antibodies and anti-cardiolipin antibodies have been shown to increase with age (42–44). Perhaps surprisingly, individuals aged between 60-74 had significantly lower ACE2-IgM than younger individuals (Supp. Fig 5). The impact of co-morbidities on anti-ACE2 antibody prevalence and levels was assessed. Conditions including hypertension, dyslipidemia, allergies, immunocompromised status, asthma, medication, cardiac conditions, and endocrine conditions were not associated with higher or lower seroprevalence of ACE2 autoantibodies (Figure 2 and Supp. Table 1). Obesity was marginally associated with a lower seroprevalence of ACE2-IgM, but this was limited to the IgM isotype. Interestingly, a higher seroprevalence of ACE2-IgG antibodies was associated with neurological conditions, but the low numbers of individuals with neurological conditions in our cohort impede further conclusions. While ACE2 has been linked with other health conditions, our cohort was sampled from the general population which limits our ability to measure associations with rare pathologies. When looking at the longitudinal trend of ACE2 autoantibodies for individuals with one or two follow-up serum collections, IgA and IgG antibodies remained relatively stable over time, whereas IgM fluctuated (Figure 3). While it is unclear what triggers the production of ACE2 autoantibodies, the increased rate of decay of IgM compared to IgG and IgA is most likely due to its intrinsic shorter half-life, and inability to bind FcRn (key receptor for IgG recycling) (45–47).

Several groups have previously reported that ACE2 autoantibodies could be induced by SARS-CoV-2 infection. For example, a small cohort study found that 81% of convalescent individuals (n=32) and 93% of hospitalized acute SARS-CoV-2 infection (n=15) had detectable ACE2 autoantibodies which were undetectable in their control group (n=13). Other groups have found similar observations (23, 24). To explore whether SARS-CoV-2 induced or correlated with ACE2 autoantibodies in our cohort, we identified two cohorts of convalescent individuals. The first cohort included individuals who had substantial levels of IgG against the N and S protein (i.e., the serological marker of a previous infection) of SARS-CoV-2 and/or declared having been infected by SARS-CoV-2 to our study coordinators. The inclusion of self-reported infection addresses concerns of misidentifying convalescent individuals in which anti-N or anti-S decayed beyond our seropositivity threshold over time. The second cohort of convalescent individuals included only individuals who had serological and/or laboratory evidence of prior infection, hereby constituting a stringent cohort eliminating subjective assessment of SARS-CoV-2 infection and patient bias. Principal component analysis failed to find clusters of convalescent individuals based on ACE2 autoantibody levels (Supp. Fig 6). In line with this, levels of IgG, IgA, and IgM ACE2 autoantibodies were not more elevated in the convalescent group (Fig 3). In fact, it appears that ACE2 IgM was elevated in non-convalescent individuals. These data demonstrate that ACE2 antibodies are not more prevalent, or present at a higher titer in SARS-CoV-2 convalescent sera. Furthermore, we asked if antibody titers against the N protein correlate with ACE2 autoantibodies. This could suggest a disease severity-dependent relationship. We found no association or association clusters between either ACE2 autoantibody isotype or SARS-CoV-2 N antibody levels (Supp. Fig 7). This further suggests that while we may not detect ACE2 autoantibody induction shortly after an infection, as shown by some groups, we show that ACE2 antibodies are decoupled with SARS-CoV-2 antibodies, and if induced in some cases, short-lived.

Our observations are in contrast with several previous studies. It is possible that high disease severity could impact ACE2 autoantibodies levels. Indeed, most studies have been focused on hospitalized or severe COVID-19 infections (22, 24). For example, a recent study found higher median ACE2 IgG intensity in moderate/severe cases of COVID-19 in contrast to mild convalescent individuals (48). However, it is important to mention that the overall seroprevalence was similar between convalescent individuals vs pre-COVID-19 sera. Other studies also found that ACE2 autoantibodies were increased in those with severe COVID-19 infection (49, 50). While severity scores for most convalescent individuals were not measured for all the serum samples, to our knowledge none had to be admitted to the ICU, suggesting mostly asymptomatic and mild-moderate COVID-19 infections. However, the link with disease severity is not obvious. In fact, in a large protein array screen of severe COVID-19 infections, ACE2 autoantibodies were only detected in one individual while several individuals had autoantibodies to numerous other proteins such as cytokines (9). Moreover, in a recent study that screened 1139 convalescent individuals for ACE2-IgG, the authors found that the majority of individuals who had ACE2-IgG did not require hospitalization following SARS-CoV-2 infection (51). It remains possible that severe infection can induce ACE2 autoantibodies. It was reported that severe SARS-CoV-2 infection can induce low selection pressure of B cells which was associated with *de novo* autoreactivity (52). In this study, we show that ACE2 autoantibodies are not exclusive to SARS-CoV-2 convalescent individuals but are common in the general population concordant with observations of these antibodies with other pathologies and in healthy populations (28, 30). These ACE2 autoantibodies may have an important regulatory role, however no data currently exist to support this hypothesis. Some studies have also suggested that ACE2 autoantibodies could arise from molecular mimicry or from spike/ACE2 aggregates making their way to germinal centers. One study noticed an increase in ACE2-IgG following vaccination (48). In the small subset of individuals vaccinated against SARS-CoV-2 in our cohort, we did not observe this effect (Supp. Fig. 8). Others have proposed anti-idiotype antibodies as a possible explanation. However, given that the seroprevalence did not differ in vaccinated or in convalescent individuals in relation to our control cohort, the data does not currently support this hypothesis. It is also possible that ACE2 autoantibodies occur as the result of molecular mimicry of another pathogen. However, there is no evidence in support.

The function, if any, of autoantibodies targeting ACE2 remains unclear. Some groups have shown data that ACE2 enzymatic activity could be inhibited by these antibodies (22), while others showed the contrary (50). We used a subset of our cohort to investigate if ACE2 enzymatic activity is impacted by the presence of plasma containing various levels of ACE2 autoantibody (Figure 4). We found that ACE2 antibodies did not impact enzymatic activity. We found that ACE2 enzymatic activity was higher in individuals with a history of previous SARS-CoV-2 infection. This could be explained by a higher amount of soluble ACE2 previously reported in convalescent individuals resulting in higher baseline enzymatic activity in our assay (53). We also explored the possibility that ACE2 autoantibodies may be protective by neutralizing SARS-CoV-2 by preventing ACE2/Spike interactions. This effect is difficult to measure in convalescent individuals given that antibodies against the spike protein would overwhelmingly contribute to the neutralization of SARS-CoV-2. As such, we depleted serum samples of antibodies that could bind to the spike protein. In some cases, the depletion process was incomplete, likely due to the initially very high titer of anti-spike antibodies. The depleted sera were then used in a protein-based neutralization assay to measure the interaction between the spike and ACE2. While some individuals still had neutralization ability, these were linked with samples with incomplete anti-spike depletion (Figure 4). In samples with no residual spike antibodies, the neutralization ability of the sera was lost, suggesting that ACE2 autoantibodies are not able to block these interactions. This suggests that either these anti-ACE2 antibodies recognize epitopes on ACE2 that do not interfere with the ACE2/spike binding or the concentration and affinity of ACE2 antibodies in sera are not able to outcompete the affinity of the spike protein for ACE2.

This study is focused on the detection of ACE2 autoantibodies in individuals who did not require hospitalization for SARS-CoV-2 infection. In addition, we were not able to longitudinally monitor ACE2 levels continuously prior to and post-SARS-CoV-2 infection as the prevalence of SARS-CoV-2 infection in the Ottawa region in late 2020 was low. Furthermore, we did not investigate the presence of ACE2 autoantibodies at mucosal surfaces, such as the upper respiratory tract. We focused our understanding of ACE2 autoantibodies in their role in SARS-CoV-2 pathogenesis, however, ACE2 autoantibodies may play a role in other physiological processes independently of infection which were beyond the scope of this work.

## Conclusion

This is the first large cohort study to report the simultaneous seroprevalence of IgG, IgA and IgM antibodies in sera able to bind ACE2. We found no evidence that ACE2 autoantibodies were induced by SARS-CoV-2 infection and were rather common in SARS-CoV-2 naïve individuals. We also demonstrate that these autoantibodies are not able to inhibit SARS-CoV-2 spike and ACE2 interactions, or able to block ACE2 enzymatic activity. The relevance of ACE2 autoantibodies in homeostasis, health, or disease remains unclear.

## Methods

### Participant recruitment and sample processing

Individuals for this study were recruited from the ongoing Stop the Spread Ottawa (SSO) study(31). Individuals 18 years and older with a history of SARS-CoV-2 infection or at risk of infection were recruited. Individuals having at least one blood draw by December 2021 with corresponding participant questionnaire were included in this study. Serum collection was performed according to current standard phlebectomy procedures and samples were de-identified prior to reception at the SARS-CoV-2 High Throughput Serology and Diagnostics Facility located at the University of Ottawa, Faculty of Medicine. Sera was stored short-term at 4°C, or frozen long-term at -80°C. Approval was granted by The Ottawa Health Science Network Research Ethics Board (Certificates: H-09-20-6135 and H-07-20-6009). Informed consent was obtained from all participants.

### SARS-CoV-2 and ACE2 antibody measurements

Antibodies against SARS-CoV-2 were measured using a high-throughput direct chemiluminescent ELISA (full methods described previously) (33). ACE2 autoantibodies were measured using a similar method on the same platform used for SARS-CoV-2 antibodies with minor modifications. Briefly, recombinant ACE2 protein (generously provided by Dr. Yves Durocher, National Research Council of Canada (NRC), Montréal) was diluted in PBS (5ug/mL working concentration) and was coated in a 384well high-binding polystyrene Nunc plate (Thermo Fisher Scientific, #460372) to a final amount of 50ng of protein per well. Coated plates were briefly centrifuged to evenly distribute ACE2 solution and incubated overnight at 4°C. Plates were then blocked for 1 h to remove non-specific binding using 80uL of 3% w/v skim milk powder dissolved in 100uL of PBS + 1% Tween (PSB-T). Samples were diluted 1 in 80 in 1% w/v skim milk powder dissolved in PBS-T, 10uL was added to respective wells, and incubated for 2h.10uL of HRP-labeled secondary antibody was added to each well and incubated for 1h.

Secondary antibodies used were anti-Human IgG#5-HRP (fusion-HRP construct) (Dr. Yves Durocher, NRC) diluted 1 in 5400; Anti-human IgA-HRP (Jackson ImmunoResearch Labs, 109-035-011) diluted 1 in 8000; and anti-human IgM-HRP (Jackson ImmunoResearch Labs, 109-035-129) diluted 1 in 9600. To detect bound secondary antibodies, 10uL of ELISA Pico Chemiluminescent Substrate (Thermo Fisher Scientific #37069) diluted 1:2 in ddH_2_O was added to each well and incubated for 5 minutes. Luminescence was measured on a Synergy NEO2 plate reader (Agilent) at 20ms/well at a read height of 1.0mm. Relative luminescence units (RLU) were blank-subtracted and used for subsequent analysis. Plate wells were washed 4x following each incubation (except the final 5 min incubation) with 100uL of PBS + 1% Tween (PSB-T) using a 405 TS/LS LHC2 plate washer (Biotek Instruments)

### ACE2 Enzymatic Activity

To investigate whether endogenous ACE2 antibodies were able to inhibit ACE2 activity, a neutralization reaction was performed. 2uL of serum samples (negative control, surveillance, convalescent, McG) were added to 0.1 ug/mL of recombinant human ACE2 (rhACE2, R&D systems, Minnesota, MN, USA, cat. 933-ZN) for 30 min at room temperature under agitation. 10uL of neutralized samples were transferred to 96-well black plates containing 15 uM of fluorogenic ACE2 substrate Mca-APK(Dnp) (Anaspec, Fremont, CA, USA, cat. AS-60757), to a final concentration of 11.25 uM (54). Samples were assayed in duplicates without ACE2 inhibitor and in singles with 10^-5^ M ACE2 inhibitor MLN-4760 (Calbiochem, San Diego, CA, cat. 530616). Following a 30-min incubation at room temperature, fluorescence was measured in a FLUOstar Galaxy fluorometer (BMG Labtech, Ortenberg, Germany), detecting emission at 405 nm and excitation at 320 nm. Data are presented as Relative Fluorescence Units (RFU) baseline-subtracted using samples with MLN-4760 and normalized by volume of input serum (RFU/uL).

### Depletion of SARS-CoV-2 spike antibodies in sera

To measure ACE2 autoantibody impact on ACE2 and SARS-CoV-2 spike interaction, serum samples were first depleted of spike-reactive antibodies. 100ng /well of spike protein (NRC) was coated in a high-binding polystyrene Nunc plate (Thermo Fisher Scientific, #460372) in a final volume of 10uL in PBS. Plates were briefly centrifuged to ensure even coating and incubated at 4°C overnight. The next day, plates were washed as described previously and blocked using 80uL of 3% w/v skim milk powder dissolved in PBST for 1 h. Plates were washed, and selected serum samples were mixed with 1:1 with 1% w/v skim milk powder dissolved in PBST (final of 50uL) and applied to the wells. The diluted samples were incubated for 2h with shaking, transferred to a new blocked plate coated with SARS-CoV-2 spike and incubated for another 2h. Samples were transferred once more and incubated overnight at 4°C to be used in the spike binding assay the next day.

### Spike and ACE2 binding assay

This protein-based assay was modified from a previously published surrogate neutralization assay(33). First, recombinant SARS-CoV-2 spike protein (generously provided by Dr. Yves Durocher, National Research Council of Canada (NRC), Montréal) was diluted in PBS and was coated in a 384 well high-binding polystyrene Nunc plate (Thermo Fisher Scientific, #460372) at 50ng/well with a final volume of 10uL/well. The plates were then centrifuged at 216xg and incubated overnight at 4°C. The plates were then washed four times with 100uL of PBS-T using a 405 TS/LS LHC2 plate washer (Biotek Instruments). The wells were then blocked with 80uL of 3% w/v skim milk powder dissolved in PBST for 1LJh and then washed (as described before). Samples that were not depleted were diluted 1 in 2 in 1% w/v skim milk powder dissolved in PBST. Previously depleted samples were already diluted 1 in 2. 20uL of each sample was added to the plate and incubated for 2h with shaking. The plates were washed once more as described previously and 6.5ng/well of recombinant biotinylated ACE2 (NRC) was added to each well in a final volume of 20uL and incubated for one hour. The plates were washed once more and bound ACE2 was detected by adding 5.2ng/well in a final volume of 20uL/well of streptavidin-peroxidase polymer (Thermo Fisher #N200), diluted in 1% w/v skim milk powder dissolved in PBST. After a 1h incubation, the plates were then washed and developed with the addition of 20uL of ELISA Pico Chemiluminescent Substrate (Thermo Fisher Scientific #37069), diluted 1:2 in MilliQ H_2_O. After a 5-minute incubation, plates were read on a Neo2 plate reader (BioTek Instruments) at 20 ms/well and a read height of 1.0 mm. Luminescence values were adjusted to the blank (serum-free, no ACE2 condition) and the average maximal signal of the plate was established using the serum-free with ACE2 condition. Percentage inhibition was then established by measuring the reduction in spike / ACE2 interaction (0% inhibition; maximal ACE2-Spike binding).

### Data analysis

Data, statistical analyses, and figures were processed in R (4.2.3). BioVenn was used to generate quantitative Venn diagrams.

## Data Availability

All data produced in the present study are available upon reasonable request to the authors

## Acknowledgements

Author contributions

YG and M-A.L designed the study. YG developed the assays and performed the experiments together with NC, PM. CC, EC, AK, recruited participants, coordinated sample collection and collected participant data. MTS and KB performed and analyzed the ACE2 enzymatic activity assay. MP, CA analyzed COVID-19 serology. YG performed the data analysis. YG and M-A.L wrote and formatted the manuscript. All authors have read, edited and approved the manuscript.

## Additional information

We acknowledge the involvement of previous and current members of the University of Ottawa Serology and Diagnostics High Throughput Facility including: Danielle Dewar-Darch, Justino Hernandez Soto, Abishek Xavier, Nicholas Bradette, Klaudia Baumann, Gwendoline Ward, Yuchu Dou and Lynda Rocheleau. We also acknowledge everyone involved in the Stop the Spread Ottawa study including participants. Production of COVID-19 reagents was financially supported by NRC’s Pandemic Response Challenge Program.

