## Supplementary Files for "Autoantibodies Targeting Angiotensin Converting Enzyme 2 Are Prevalent and Not Induced by SARS-CoV-2 Infection"

*Corresponding author

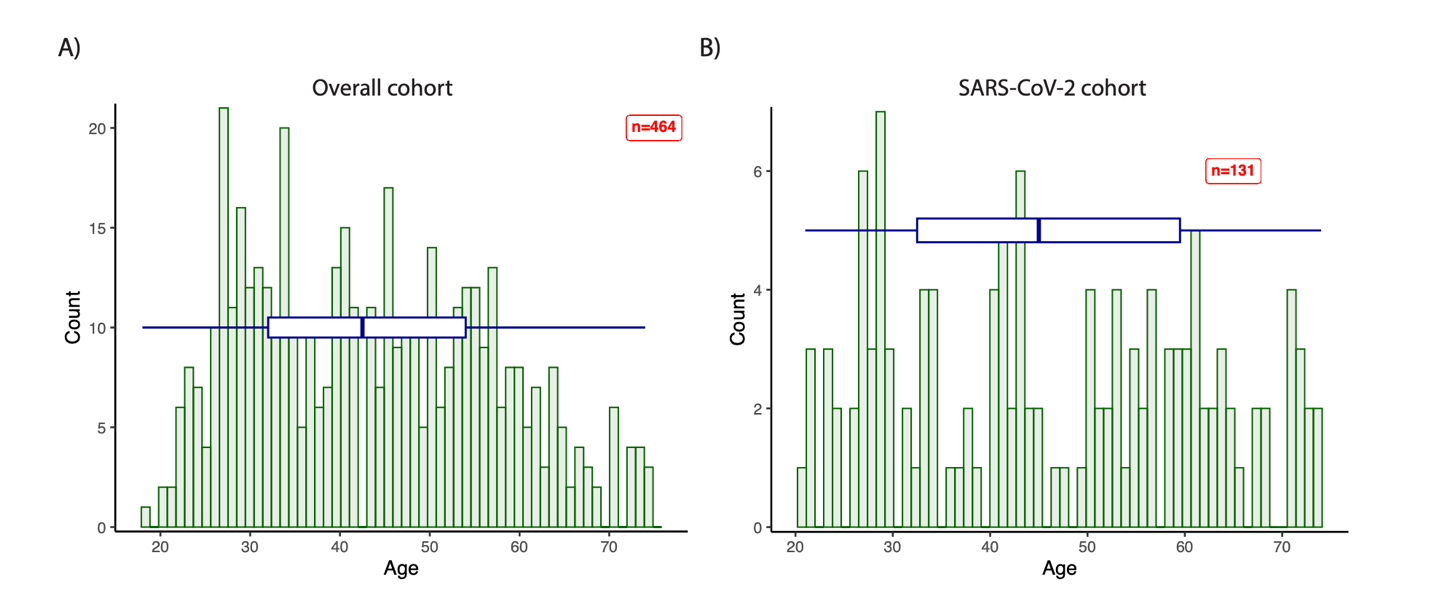

**Supplemental Figure 1. Cohort demographics.** A) Age (years) distribution in the whole cross-sectional cohort (n=464). B) Age (years) distribution of individuals with serological evidence of prior SARS-CoV-2 infection and or self-reported a prior infection (n=131). The number of individuals is represented as count on the y axis of the graphs. The boxplots overlayed on the histogram represent the distribution of the cohort, with the median, and different quantiles.

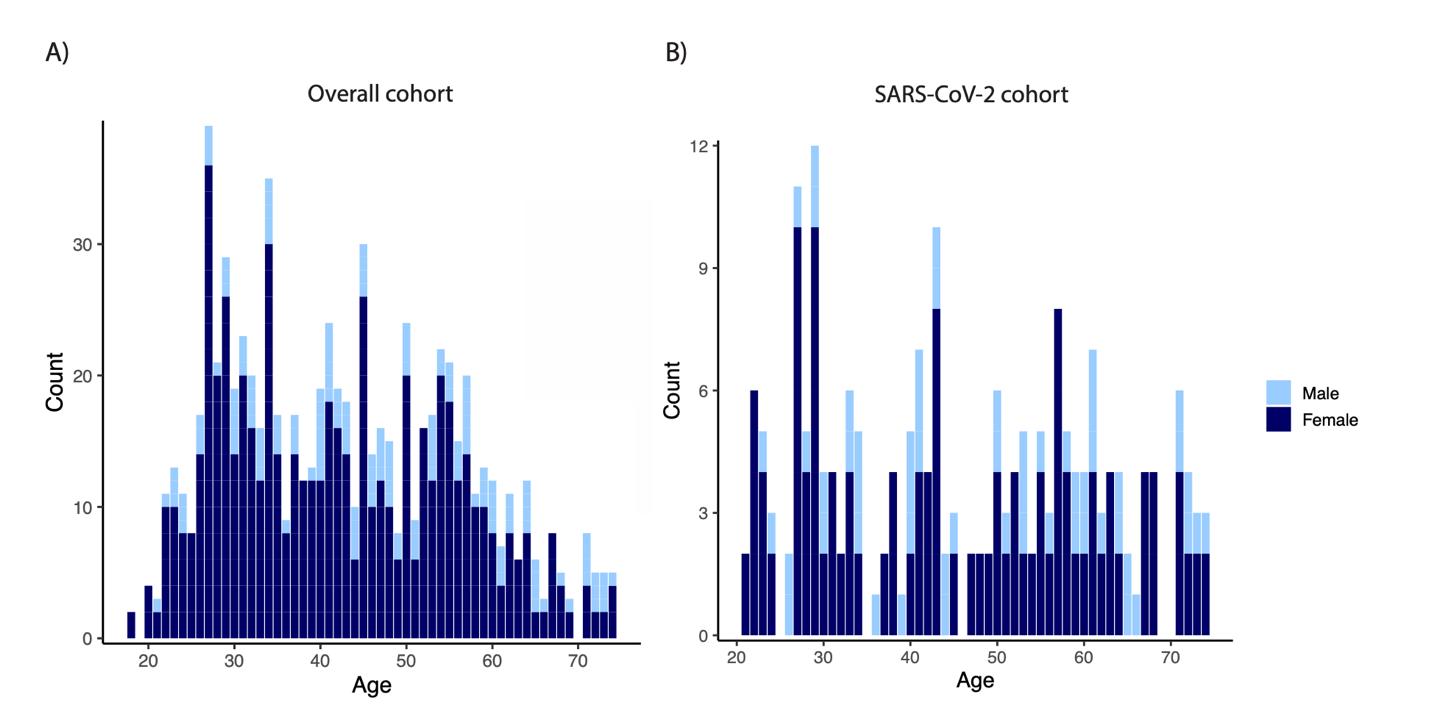

**Supplemental Figure 2. Cohort demographics by sex.** A) Age (years) and sex distribution in the whole cross-sectional cohort (n=464). B) Age (years) and sex distribution of individuals with serological evidence of prior SARS-CoV-2 infection and or self-reported a prior infection (n=131). The number of individuals is represented as count on the y axis of the graphs.

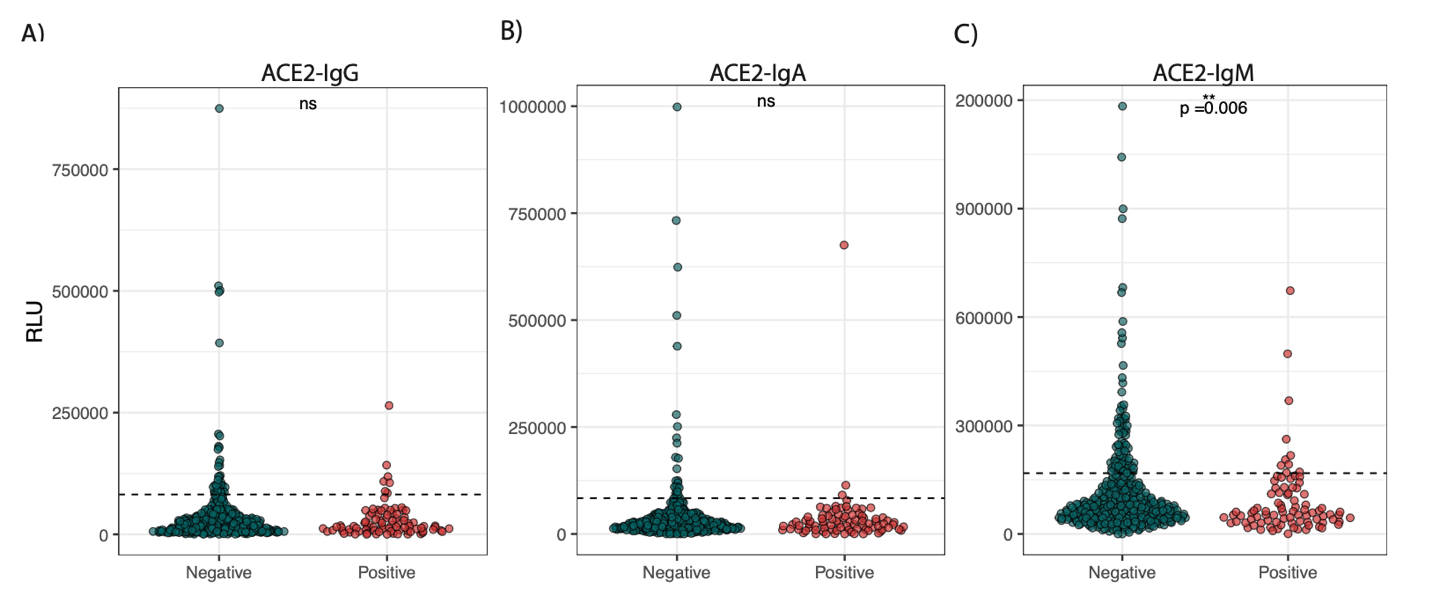

**Supplemental Figure 3. ACE2 autoantibodies levels according to serological evidence of SARS-CoV-2 infection.** A-C) IgG, IgA, IgM ACE2 autoantibodies levels are shown for individuals who never had SARS-CoV-2 and for individuals who had serological markers of previous infection (anti-SARS-CoV-2 N and S IgG positive). Unpaired two-sided Wilcoxon test was used to establish statistical significance (n=464), * p< 0.05, ** p< 0.01, ***p<0.001, ****p<0.0001.

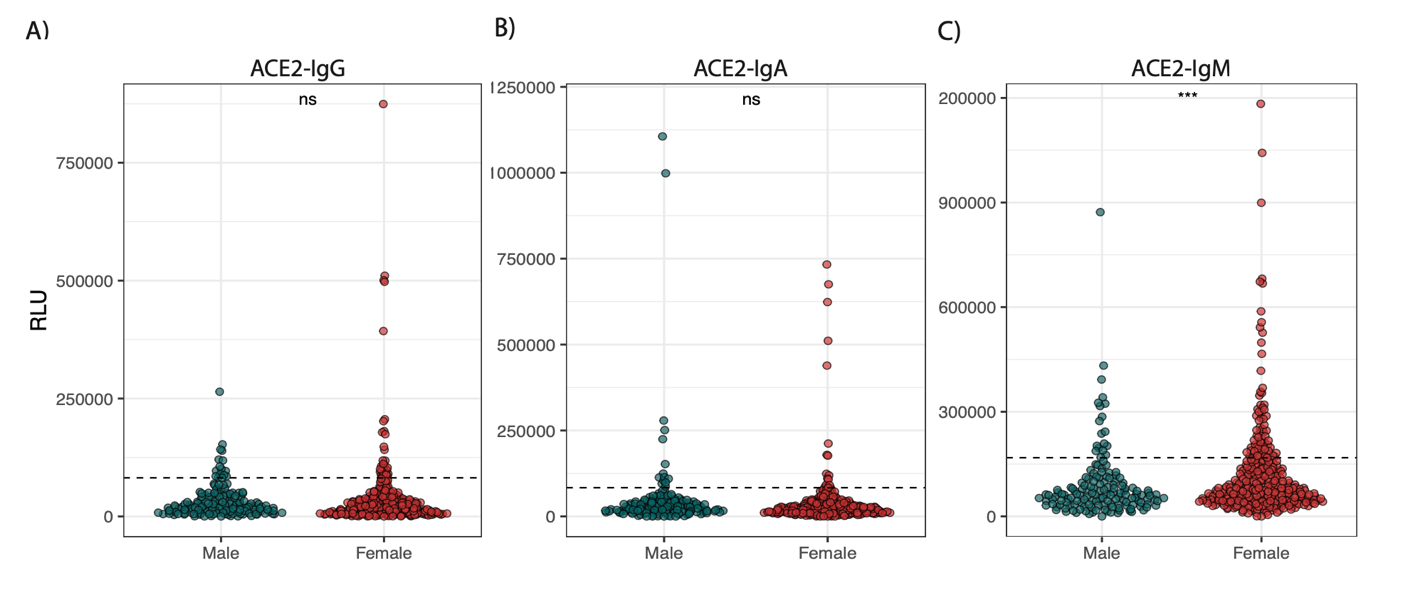

**Supplemental Figure 4. ACE2 autoantibodies levels by sex and by isotype.** A-C) IgG, IgA, IgM ACE2 autoantibodies levels are shown for all individuals in the cross-sectional cohort. Unpaired two-sided Wilcoxon test was used to establish statistical significance (n=464), * p< 0.05, ** p< 0.01, ***p<0.001, ****p<0.0001.

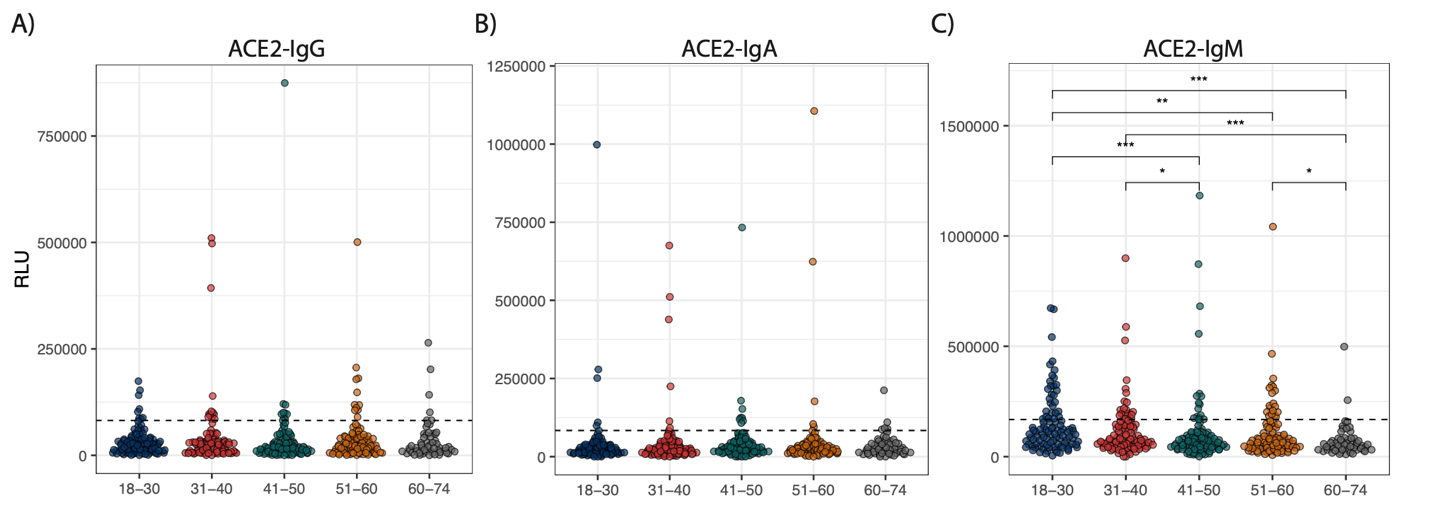

**Supplemental Figure 5. ACE2 autoantibodies levels by age groups and by isotype.** A-C) IgG, IgA, IgM ACE2 autoantibodies levels are shown for all individuals in the cross-sectional cohort. Kruskal-Wallis test was used for global statistical significance and Wilcoxon test was used for multiple comparisons for pairwise differences (n=464), * p< 0.05, ** p< 0.01, ***p<0.001, ****p<0.0001.

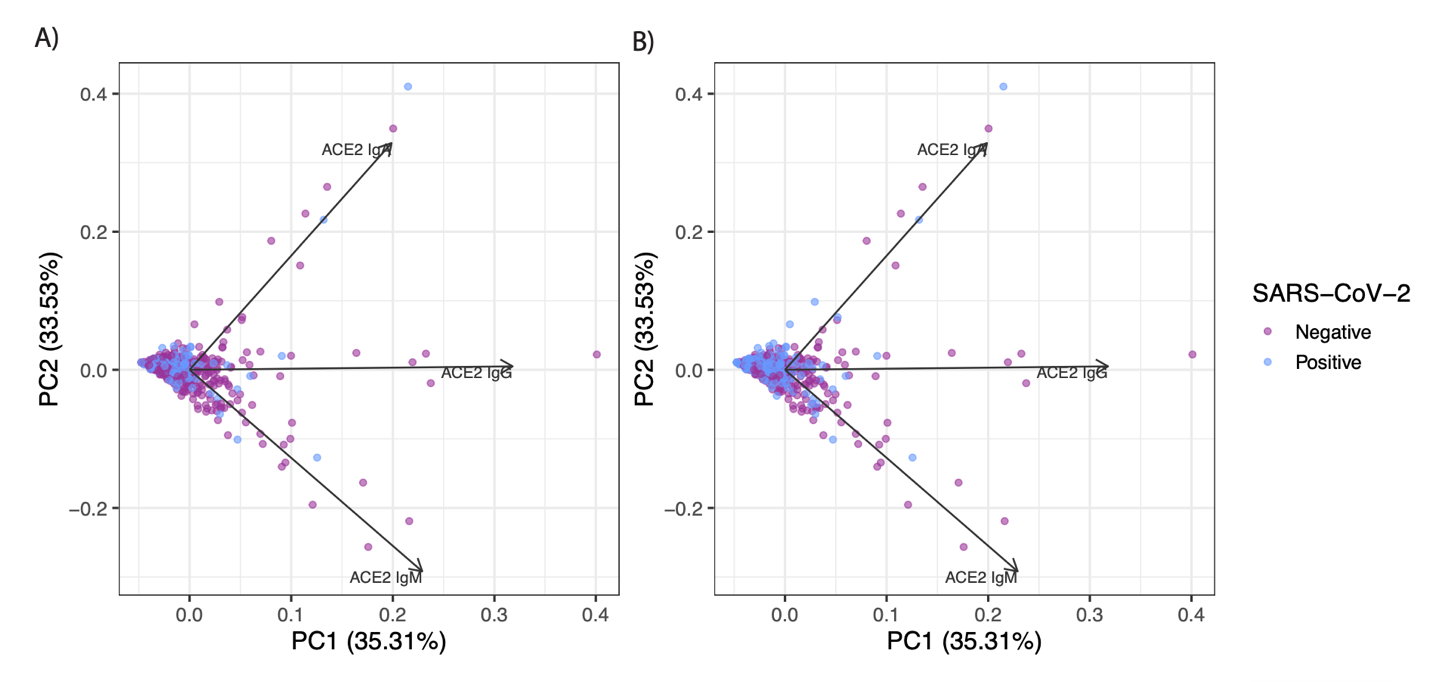

**Supplemental Figure 6. Principal component analysis of ACE2 autoantibodies and classified by prior SARS-CoV-2 infection status.** Principal component analysis (PCA) showing the clustering between SARS-CoV-2 prior infection and ACE2 autoantibody (IgG, IgA, IgM) levels. A) Individuals who have serological markers of SARS-CoV-2 infection. B) Individuals having serological markers of prior SARS-CoV-2 infection and/or self-reported a prior infection.

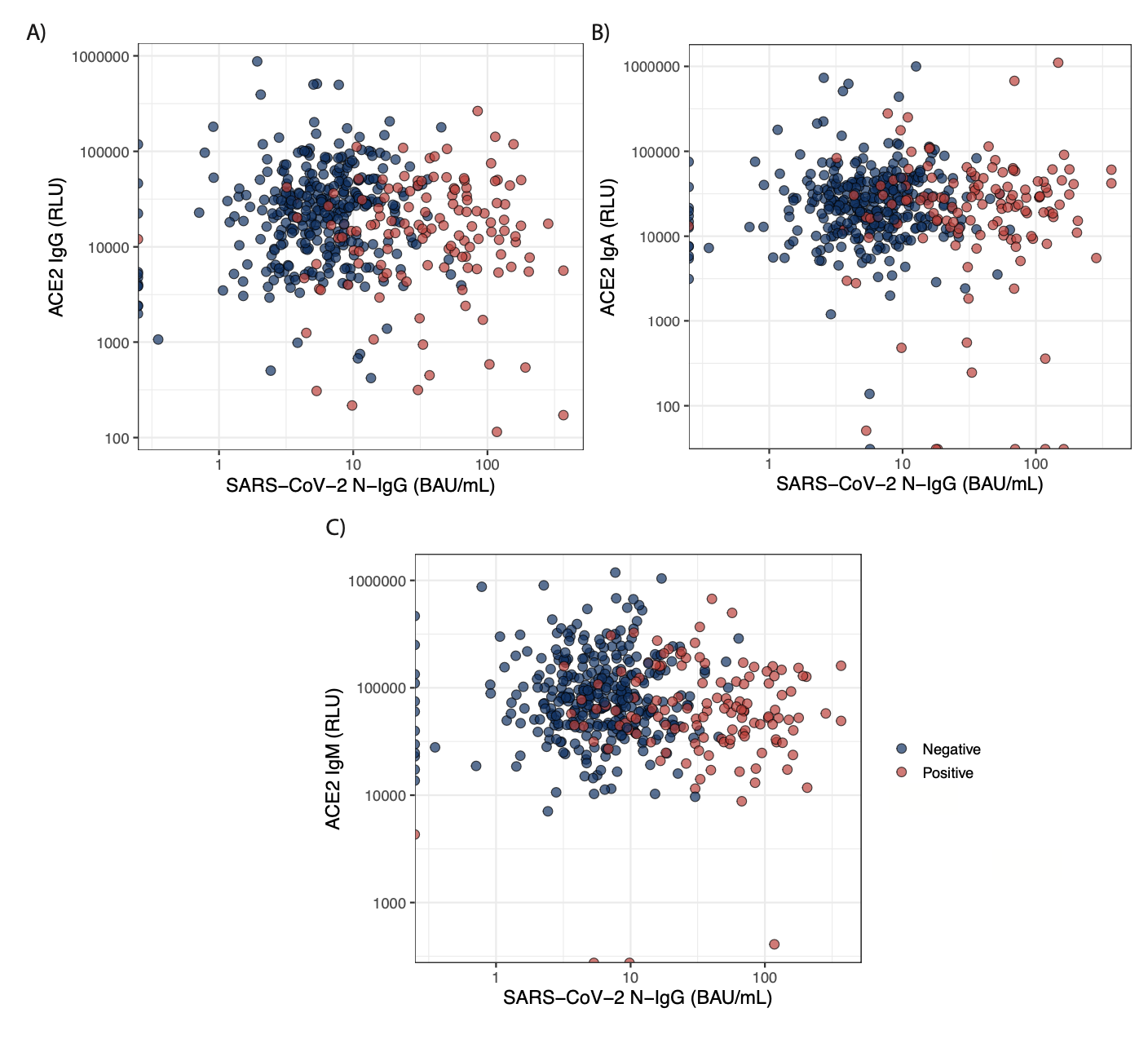

**Supplemental Figure 7. Correlation between ACE2 autoantibodies and SARS-CoV-2 nucleocapsid IgG antibodies.** Individuals were further classified on their SARS-CoV-2 infection status (identified by serology or self-reported). Correlation between A) ACE2-IgG B) ACE2-IgA C) ACE2-IgM antibodies and N-IgG. X and Y axis were formatted in Log10 scales.

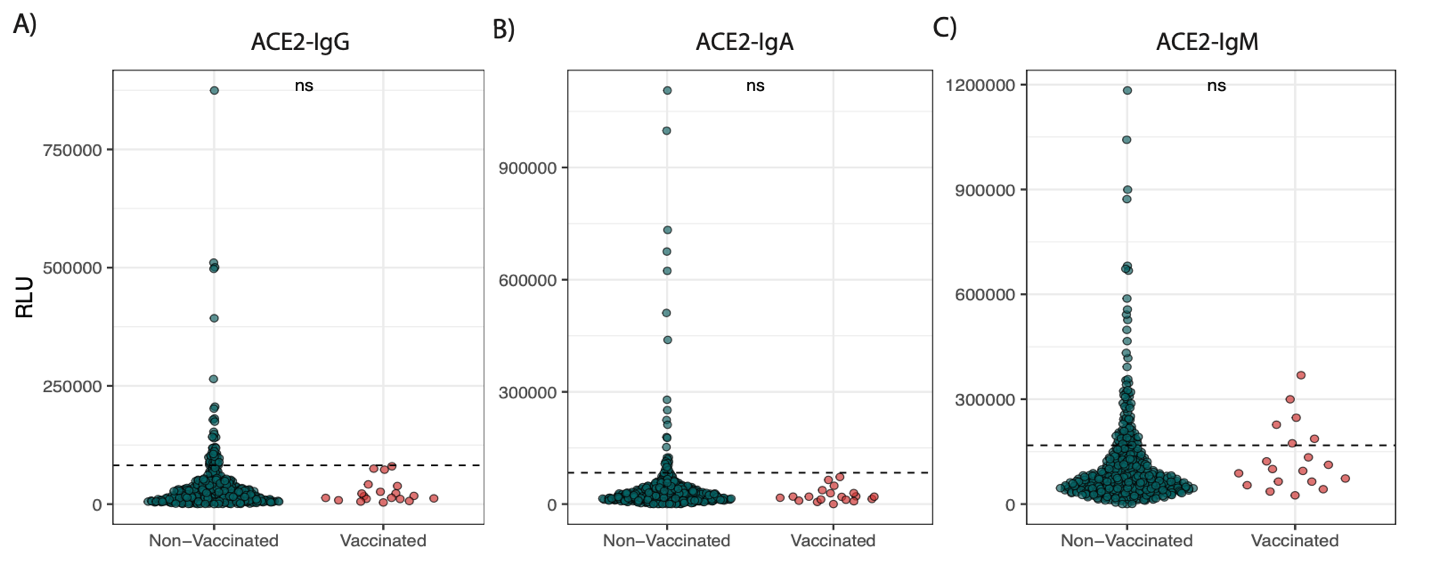

**Supplemental Figure 8. ACE2 autoantibodies levels by vaccination status.** A-C) IgG, IgA, IgM ACE2 autoantibodies levels are shown for all individuals in the cross-sectional cohort. Unpaired two-sided Wilcoxon test was used to establish statistical significance (n=464), * p< 0.05, ** p< 0.01, ***p<0.001, ****p<0.0001.

**Supplemental Table 1. ACE2 autoantibody seroprevalence odd ratios and 95% confidence interval by isotype.** Demographic and clinical information collected as part of the study were evaluated on their influenced on ACE2 autoantibody seroprevalence (IgG, IgA, IgM).

|  | ACE2 IgG (OR, (95%CI) | ACE2 IgA (OR, (95%CI) | ACE2 IgM (OR, (95%CI) |
| --- | --- | --- | --- |
| Female | 0.97 (0.52 - 1.83) | 0.50 (0.23 - 1.06) | 1.95 (1.12 - 3.38) |
| Male | 1.03 (0.55 - 1.94) | 2.01 (0.94 - 4.28) | 0.51 (0.30 - 0.89) |
| SARS-CoV-2 infection | 0.48 (0.22 - 1.05) | 1.37 (0.62 - 3.02) | 0.51 (0.29 - 0.92) |
| Immunocompromised | 2.01 (0.65 - 6.21) | - * | 0.96 (0.32 - 2.91) |
| Obesity | 1.51 (0.69 - 3.28) | 1.28 (0.47 - 3.50) | 0.38 (0.16 - 0.94) |
| Hypertension | 0.56 (0.17 - 1.89) | 1.03 (0.30 - 3.53) | 0.61 (0.25 - 1.48) |
| Dyslipidemia | 0.68 (0.20 - 2.30) | 1.77 (0.58 - 5.38) | 0.91 (0.39 - 2.14) |
| Asthma | 0.75 (0.26 - 2.18) | 1.39 (0.46 - 4.16) | 0.97 (0.45 - 2.09) |
| Allergies | 1.48 (0.82 - 2.70) | 1.07 (0.50 - 2.28) | 1.08 (0.68 - 1.73) |
| Taking medication | 0.89 (0.47 - 1.68) | 1.00 (0.44 - 2.25) | 1.14 (0.69 - 1.91) |
| Neurological conditions | 5.48 (1.27 - 23.69) | 2.18 (0.26 - 18.37) | 1.45 (0.29 - 7.33) |
| Cardiac conditions | 3.35 (0.86 - 13.08) | - * | 0.43 (0.05 - 3.37) |
| Endocrine conditions | 1.71 (0.68 - 4.34) | 0.82 (0.19 - 3.59) | 0.64 (0.24 - 1.68) |

*Odd ratios and confidence intervals could not be measured given the low seroprevalence of IgA anti-ACE2 in these groups

**Supplemental Table 2. Participant demographic information for enzymatic assay cohort.** The enzymatic activity of ACE2 was measured in the presence of serum from a subset of individuals of the initial cohort.

|  | **Enzymatic Assay cohort**  (n=103) |
| --- | --- |
| **Age (y.)** |  |
| Mean (SD) | 45.73(14.0) |
| Median | 45.0 |
| Range | 24-73 |
| **Sex** |  |
| Female Nb. (%) | 65(63.1) |
| **Previous SARS-CoV-2 infection Nb. (%)**  (Serology and/or self-reported) | 38(36.9) |
| **ACE2 seroprevalence** |  |
| IgG (%) | 9.71 |
| IgA (%) | 7.77 |
| IgM (%) | 15.5 |

**Supplemental Table 3. Participant demographic for the ACE2 and Spike attachment.** The ability of ACE2 to bind the spike protein of SARS-CoV-2 was measured in the presence of serum from a subset of individuals of the initial cohort.

|  | **Combined**  (n=55) |
| --- | --- |
| **Age (y.)** |  |
| Mean (SD) | 46.4(15.7) |
| Median | 45.0 |
| Range | 25-73 |
| **Sex** |  |
| Female Nb. (%) | 27(49.1) |
| **Previous SARS-CoV-2 infection Nb. (%)**  (Serology and/or self-reported) | 36(65.5) |
| **ACE2 seroprevalence** |  |
| IgG (%) | 9.09 |
| IgA (%) | 5.45 |
| IgM (%) | 16.4 |
